# A 5-year Review of Incidence, Presentation and Management of Bartholin Gland Cysts and Abscesses in a Tertiary Hospital, Yenagoa, South-South Nigeria

**DOI:** 10.1101/2022.05.01.22274551

**Authors:** PC Oriji, DO Allagoa, AE Ubom, VK Oriji

**Author notes:** **Correspondence:** Dr. Dennis Oju Allagoa.

## Abstract

**Background:** Bartholin gland cysts and abscesses are common in women of reproductive age and declines after menopause. Organisms implicated in Bartholin abscess include, *Neisseria gonorrhoeae, Chlamydia trachomatis, Escherichia coli, Staphylococcus aureus* and Bacterioides spp.

**Aim:** To determine the incidence, presentation and management of Bartholin gland cysts and abscesses at the Federal Medical Centre, Yenagoa, Bayelsa State, South-South Nigeria, over a five-year period.

**Settings and Design:** This retrospective study was conducted in the Gynaecological Unit of the Federal Medical Centre, Yenagoa, Bayelsa State, South-South, Nigeria, between January 1, 2016 and December 31, 2020.

**Materials and Methods:** Relevant data were retrieved, entered into a pre-designed proforma, and analysed.

**Statistical Analysis:** Statistical Package for the Social Sciences version 25.0 was used for data analysis. Results were presented in frequencies and percentages for categorical variables.

**Results:** There were 2,478 gynaecological cases managed in our Centre; out of which there were 26 cases of Bartholin cyst and abscess, giving an incidence of 1.05%. Most of the women were ≤ 30 years (14, 53.8%), single (17, 65.4%), nulliparous (13, 50.0%), traders (11, 42.3%), with only primary/secondary education (18, 69.2%). The left Bartholin gland was the most frequently affected (17, 65.4%). A positive microbial culture was obtained in 84% of cases, with *Staphylococcus aureus* and *Escherichia coli* being the isolated organisms. Marsupialisation was the treatment modality in all the patients.

**Conclusion:** Women of reproductive age-group should be counselled on this condition and encouraged to keep good perineal hygiene and better sexual conduct so as to reduce the risk of Bartholin cysts and abscesses.

## INTRODUCTION

The Danish anatomist, Caspar Bartholin was the first to describe the gland in 1677, hence the name Bartholin gland.^1^ The Bartholin gland is also called the greater vestibular gland. Each Bartholin gland is approximately 0.5 cm in size and drains drops of mucous into a duct 2.5 cm long.^1–3^ There are two Bartholin glands, each found in the labia minora at the 4– and 8– o’ clock positions respectively on each side of the vaginal orifice, just below the hymenal ring.^2,3^ The organs play an important role in the female reproductive system and its main function is to secrete mucus that lubricates the vagina and vulva especially during sexual intercourse.^3^ The glands are generally not palpable, apart from when they are diseased.^2,3^ Bartholin cyst form when the ostium of the Bartholin duct gets occluded with resultant accumulation of mucous secretion from the gland, while Bartholin abscess results from either primary gland infection or infection of a Bartholin cyst.^4^

Bartholin gland cysts and abscesses are common in women of reproductive age and declines after menopause.^5^ A previous Bartholin cyst/abscess is a risk factor for a repeat Bartholin cyst/abscess.^6^ Bartholin abscess occurs in about 2% of women.^2^ The causes of duct blockage include vestibular injury, iatrogenic occlusion from stitches placed during surgery, inflammation from infection, congenital narrowing of the duct and inspissation of mucus leading to plugging.^2^ Bartholin abscess is usually polymicrobial and occasionally attributable to sexually transmitted pathogens.^2,6^ Organisms implicated in Bartholin abscess include, *Neisseria gonorrhoeae, Chlamydia trachomatis, Escherichia coli, Staphylococcus aureus* and Bacterioides spp.^2,6^

Bartholin cysts are usually painless, asymptomatic and often unilateral. They may get enlarged and cause discomfort, especially during sitting, ambulating and sexual intercourse.^2^ Bartholin abscess commonly presents with excruciating, progressive vulva pain that limits movement, and in some instances, alters the patient’s gait.^2^ The differential diagnoses include sebaceous cyst, fibroma, lipoma, hydradenoma, haematoma, vulval varicosity, inclusion cyst, endometriotic cyst, cystadenoma of sweat glands, Gartner’s duct cyst and adenocarcinoma of the Bartholin glands, especially in postmenopausal women.^7–9^

The principle of management of Bartholin abscess includes adequate analgesia, surgical drainage, bacteriological investigation of recovered specimen and appropriate antibiotic therapy. The definitive management of Bartholin cyst and abscess is surgical. They include aspiration, simple incision and drainage, puncture of the cyst or abscess and placement of word catheter (performed as a simple office procedure), marsupialisation, gland excision, as well as cauterisation with carbon dioxide laser and silver nitrate.^10^ The main disadvantage of aspiration and incision and drainage is the high recurrence rate associated with them.^2^ Excision of the gland, which is indicated for recurrent cases is associated with significant haemorrhage, haematoma formation, postoperative pain, infection and dyspareunia from vaginal dryness and scarring.^11^ Modern methods are being discussed, including the use of hydrodissection for excision, as well as magnetic resonance imaging in devising treatment for recurrent cysts.^10^

Marsupialisation is the most common treatment modality. It is a cheap, simple-technique procedure that is associated with minimal blood loss and preserves the glandular function.^4^ It is however, associated with a recurrence rate of up to 10%.^12^ The different laser surgical modalities of treatment have the advantage of faster wound healing, office procedure pattern, and minimal scar tissue formation.^11^ They are however, associated with a higher recurrence rate than marsupialisation, expensive and require more skill.^11^ The recent trend towards the placement of Word catheter, has transformed the treatment of Bartholin cyst/abscess, and has made the management of these cases an office event.^13^ The Word catheter is not readily available in our Centre and as a result yet to be used in the management of our patients. The objective of this study is to determine the incidence, presentation and management of Bartholin gland cysts and abscesses at the Federal Medical Centre, Yenagoa, Bayelsa State, South-South Nigeria.

## MATERIALS AND METHODS

This retrospective study was conducted in the Gynaecological Unit of the Federal Medical Centre, Yenagoa, Bayelsa State, South-South, Nigeria, between January 1, 2016 and December 31, 2020. All the women managed for Bartholin cysts and abscesses in our facility during the period under review, were included in this study.

Relevant data were retrieved from the case records of the women using a purpose-designed proforma. These data included sociodemographic characteristics, clinical characteristics and management of the patients.

Data extracted was analysed using Statistical Package for the Social Sciences version 25.0. Results were presented in frequencies and percentages for categorical variables.

## RESULTS

There were 2,478 gynaecological cases managed in our Centre; out of which there were 26 cases of Bartholin cyst and abscess, giving an incidence of 1.05%.

### Sociodemographic characteristics

Most of the women were ≤ 30 years (14, 53.8%), single (17, 65.4%), nulliparous (13, 50.0%), traders (11, 42.3%), with only primary/secondary education (18, 69.2%), as seen in Table 1.

**Table 1:** Sociodemographic characteristics.

| Characteristics | Frequency, $n = 26$ | Percentage (%) |
| --- | --- | --- |
| **Age Group (years)** |  |  |
| 21 – 25 | 8 | 30.8 |
| 26 – 30 | 6 | 23.1 |
| 31 – 35 | 10 | 38.4 |
| > 35 | 2 | 7.7 |
| **Marital Status** |  |  |
| Single | 17 | 65.4 |
| Married | 9 | 34.6 |
| **Level of Education** |  |  |
| Primary | 1 | 3.8 |
| Secondary | 17 | 65.4 |
| Tertiary | 8 | 30.8 |
| **Occupation** |  |  |
| Trader | 11 | 42.3 |
| Civil servant | 8 | 30.8 |
| Unemployed | 6 | 23.1 |
| Professional | 1 | 3.8 |
| **Parity** |  |  |
| Nulliparity | 13 | 50.0 |
| Primiparity | 5 | 19.2 |
| Multiparity | 8 | 30.8 |

### Clinical characteristics, microbiology, and management

Bartholin abscess was the predominant presenting pathology (21, 80.8%), with vulval swelling being the most common presenting symptom, seen in all the patients. The left Bartholin gland was the most frequently affected (17, 65.4%), and multiple sexual partners was the commonest predisposing factor, seen in almost one-half of the patients (12, 46.2%). A positive microbial culture was obtained in 84% of cases, with *Staphylococcus aureus* and *Escherichia coli* being the isolated organisms. Marsupialisation was the treatment modality in all the patients, and this was done under local infiltration in more than three-fourth (21, 80.0%) of the patients. These characteristics are depicted in Table 2 and 3, and Figure 1.

**Table 2:** Clinical characteristics and predisposing factors to Bartholin cyst and abscess.

| Characteristics | Frequency, *n* = 26 | Percentage (%) |
| --- | --- | --- |
| **Clinical presentation\*** |  |  |
| Vulva swelling | 26 | 100.0 |
| Vulva pain | 23 | 88.5 |
| Dyspareunia | 18 | 69.2 |
| Walking difficulty | 8 | 30.8 |
| **Site of disease** |  |  |
| Left | 17 | 65.4 |
| Right | 8 | 30.8 |
| Bilateral | 1 | 3.8 |
| **Type of disease** |  |  |
| Abscess | 21 | 80.8 |
| Cyst | 5 | 19.2 |
| **Predisposing factors\*** |  |  |
| Multiple sexual partners | 12 | 46.2 |
| Previous cyst/abscess | 8 | 30.8 |
| Sexually transmitted infections | 7 | 26.9 |
| None identified | 5 | 19.2 |

*\*More than one clinical presentation/predisposing factor applies*

**Table 3:**
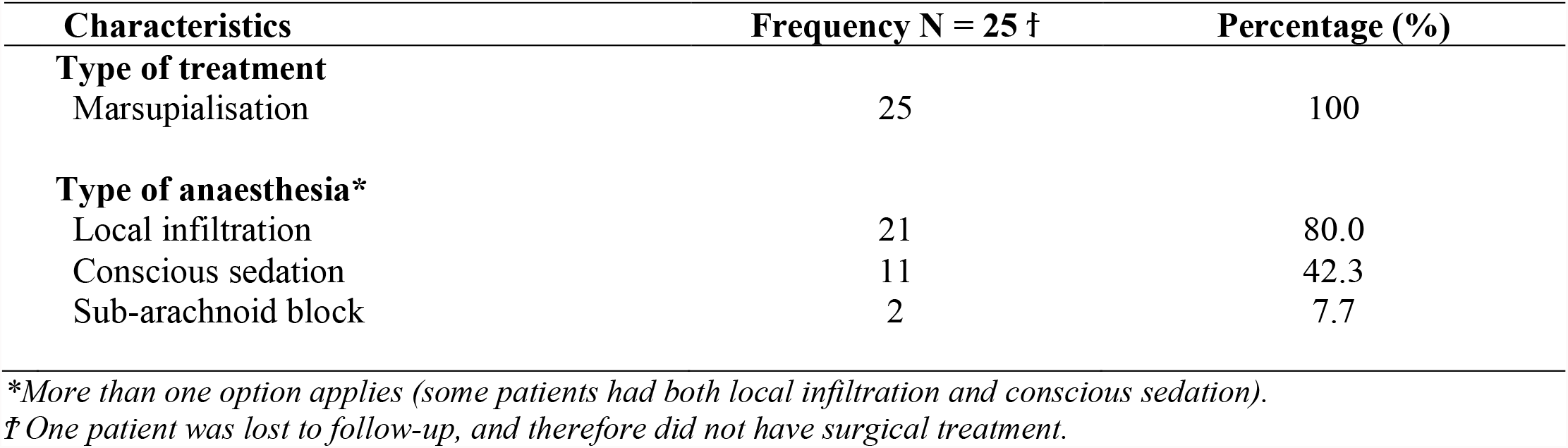
Treatment and type of anaesthesia.

**Figure 1:**
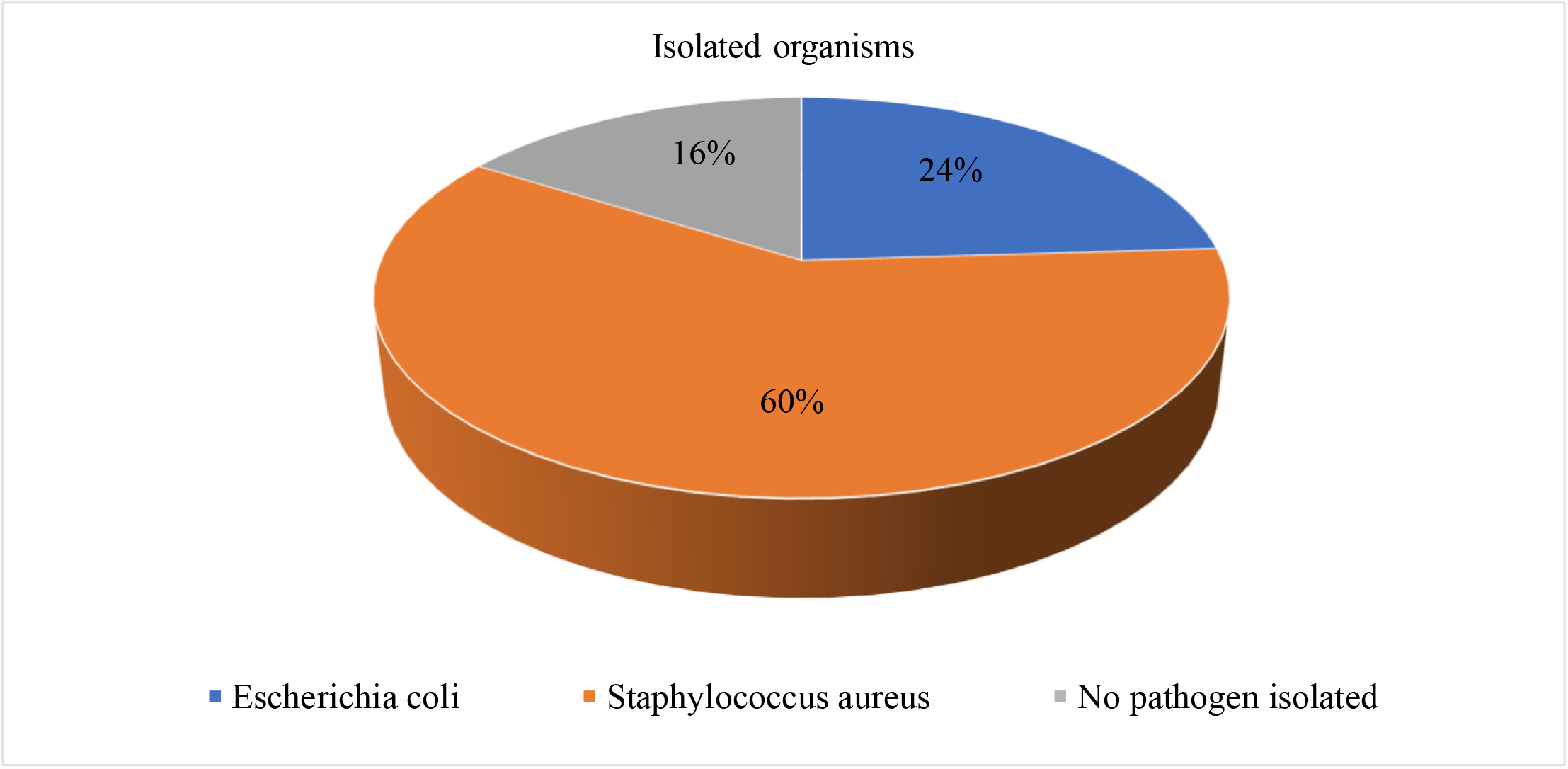
Isolated organisms from the swab culture.

## DISCUSSION

The predominantly reproductive age-group, single, nulliparous, and low socioeconomic status demographics of our study population has been corroborated by other authors.^1,2^ More than 50% of the women in our study were between 21 – 30 years of age. Bartholin cyst and abscess typically affect sexually active women between the ages of 20 – 30 years, with a lifetime risk of 2%, which decreases after menopause.^1,6,^ This is so because beyond 30 years of age, the Bartholin glands gradually begin to involute.^14^ More so, in the pre-menopausal period, mucus production by the glands is higher, and the secretions, more viscous, increasing the tendency for plugging of the ducts.^5^ After menopause however, hypoestrogenism causes atrophy and less mucus production by the Bartholin glands. The tendency for plugging of the gland ducts, and the consequent formation of cysts and abscesses therefore reduces following menopause.^5^ When Bartholin cysts or abscesses occur in postmenopausal women, malignancy should be suspected.^15^ Both conditions also occur very rarely in prepubertal females, as the Bartholin glands begin to function at puberty.^16^ Bartholin cyst and abscess occur more frequently in sexually active women because oedema from friction during sexual intercourse makes the Bartholin glands prone to obstruction at their orifices into the vestibule.^17^

Bartholin abscesses are three times more common than cysts.^3^ This was similarly the finding in our study, as 80% of our patients presented with an abscess. The predominance of Bartholin gland abscesses could be because Bartholin cysts are mostly asymptomatic, and therefore, usually ignored by the majority of patients, until the cyst becomes infected and symptomatic.^18^ As corroborated by our study findings, Bartholin cyst and abscess typically present as unilateral, tender, and inflamed posterior labial mass.^19^ The left gland was the most frequently affected in our study. This was similarly the finding of John et al.^1^ Berger et al however, found more cysts on the right side.^15^ Occasionally, affectation could be bilateral.^1^ This was the case in less than 5% of our patients. Vulvar pain, reported by nearly 90% of our patients, results from pressure of the entrapped secretion within the occluded gland duct. The pain is worsened with further mucus production and movement during sexual intercourse, resulting in dyspareunia,^20^ which was the presenting symptom in nearly 70% of our patients. As reported by 30% of the women in our study, vulvar pain may be severe enough to restrict walking and other physical activities.^2^

The most common predisposing factor, reported by almost one-half of the women in this study, was multiple sexual partners. The association of Bartholin cyst and abscess with sexual activity plausibly explains this finding. Previous history of Bartholin cyst/abscess was the risk factor in 30% of the patients. Recurrence rate of Bartholin cyst and abscess is as high as 38%.^16^ Sexually transmitted infections were the least commonly reported predisposing factor in our study, as STIs play a minor role in the aetiology of Bartholin cyst and abscess.^17^ As the vulvar is colonized by organisms commonly found on the skin, vagina, and rectum, microbes causing Bartholin gland abscesses are usually polymicrobial, with *Staphylococcus aureus and Escherichia coli*, which were the two isolates in all cases with a positive microbial culture in our study, being the most predominant organisms.^14,21^ Uncommonly, *Neisseria gonorrhoea* and *Chlamydia trachomatis* may also be implicated.^22^ Hence, a full STI screen is recommended in women presenting with Bartholin gland cysts and abscesses.^23^ *Neisseria gonorrhoea* and *Chlamydia trachomatis* are not easily isolated by laboratory culture tests. This may be a plausible reason for their low incidence in swab culture tests, and in this study. Nucleic acid amplification test (NAAT) is the gold standard for detection of *Neisseria gonorrhoea* and *Chlamydia trachomatis*.^24^ It is highly sensitive and specific in detecting the presence of these organisms, even in asymptomatic patients.^25^

Surgery is the mainstay of management of Bartholin cyst/abscess, and marsupialization and fistulisation are the most common treatment modalities.^14^ Marsupialization, which involves wide excision, drainage and eversion of the cyst mucosa to the vaginal skin, is favoured in resource-poor settings like ours, because it is cheap, fast, effective, and can be performed as an outpatient procedure.^1,26^ Hence, all the patients in our study had marsupialisation done. Marsupialisation is also less haemorrhagic, and preserves the function of the Bartholin gland,^4^ as it creates a new outflow tract (consisting of the cyst wall), following shrinkage and re-epithelization of the cyst wall over time.^12^ It can be performed under general anaesthesia, local anaesthesia, or pudendal nerve block.^10,12^ In our study, local infiltration was used in 80% of the cases. The healing and recurrence rate of 3 – 10% of marsupialization is similar to fistulisation, silver nitrate and alcohol sclerotherapy.^12,14,26^ Needle aspiration and incision and drainage, which are the simplest surgical treatment modalities, are not recommended owing to high recurrence rates.^14,17^

## LIMITATION

This is a single hospital-based study, and therefore the findings cannot be used to draw general conclusions.

## CONCLUSION

Women of reproductive age should be counselled on this condition and encouraged to keep good perineal hygiene and better sexual conduct so as to reduce the risk of Bartholin cyst and abscess.

## Data Availability

All data produced in the present work are contained in the manuscript.

## ACKNOWLEDGEMENT

The authors appreciate the Head of Department and members of staff of our Institution’s Medical Records Department for releasing the patients’ folders for this study.

## SOURCE OF FUNDING

The research was funded by the authors.

## CONFLIT OF INTEREST

The authors declare that there are no conflicts of interest.

## AUTHORS’ CONTRIBUTIONS

PCO conceptualised the study, collected and collated data, and wrote the introduction and results. DOA designed the study, wrote the first draft of the manuscript and supervised the entire research. AEU wrote the discussion. VKO carried out microbiological studies. All authors read and approved the final manuscript.

## ETHICAL APPROVAL

The research work was examined and approved by the hospital research and ethics committee.

## Notes

### Competing Interest Statement

The authors have declared no competing interest.

### Funding Statement

This study was funded by the authors

### Author Declarations

Federal Medical Centre, Yenagoa, Bayelsa State, South-South Nigeria. Research and Ethics Committee.

